# Prevalence and risk factors of adverse effects after the first COVID-19 booster dose: evidence from Greece

**DOI:** 10.1101/2023.03.27.23287816

**Authors:** Petros Galanis, Aglaia Katsiroumpa, Irene Vraka, Vanessa Chrysagi, Olga Siskou, Olympia Konstantakopoulou, Theodoros Katsoulas, Parisis Gallos, Daphne Kaitelidou

**Affiliations:** Clinical Epidemiology Laboratory, Faculty of Nursing, National and Kapodistrian University of Athens, Athens, Greece; Department of Radiology, P. & A. Kyriakou Children’s Hospital, Athens, Greece; Department of Tourism Studies, University of Piraeus, Piraeus, Greece; Center for Health Services Management and Evaluation, Faculty of Nursing, National and Kapodistrian University of Athens, Athens, Greece; Faculty of Nursing, National and Kapodistrian University of Athens, Athens, Greece

**Keywords:** adverse effects, risk factors, COVID-19, vaccines, vaccination, prevalence

## Abstract

**Background:** In general, COVID-19 vaccines are safe and effective, but minor adverse effects are common.

**Objective:** To estimate the prevalence of adverse effects after the first COVID-19 booster dose, and to identify possible risk factors.

**Material and methods:** We conducted a cross-sectional study with a convenience sample in Greece during November 2022. We measured several adverse effects after the booster dose, such as pain at the injection site, swelling at the injection site, fatigue, muscle pain, headaches, fever, chills, nausea, etc. We considered gender, age, chronic disease, self-assessment of health status, COVID-19 diagnosis, and self-assessment of COVID-19 course as possible predictors of adverse effects.

**Results:** In our sample, 96% developed at least one adverse effect. Half of the participants (50.2%) developed one to five adverse effects, 35.9% developed six to ten adverse effects, and 9.5% developed 11 to 16 adverse effects. Mean number of adverse effects was 5.5. The most frequent adverse effects were pain at the injection site (84.3%), fatigue (70.8%), muscle pain (61%), swelling at the injection site (55.2%), headache (49.8%), fever (42.9%), and chills (41%). Females developed more adverse effects than males (p<0.001). Also, we found a positive relationship between severity of COVID-19 symptoms and adverse effects of COVID-19 vaccines (p=0.005). Moreover, younger age was associated with increased adverse effects (p<0.001).

**Conclusions:** Almost all participants in our study developed minor adverse effects after the booster dose. Female gender, worse clinical course of COVID-19, and decreased age were associated with increased adverse effects.

## Introduction

Development of safe and effective vaccines against COVID-19 have offered protection against SARS-CoV-2 infections and have significantly reduced hospitalizations, intensive care unit admissions, and deaths [1,2]. However, the effectiveness and efficiency of COVID-19 vaccines decrease significantly six months after full vaccination [3]. For this reason, many countries have adopted booster doses after the initial full vaccination to boost the immune system of the population. Indeed, the effectiveness of the first booster dose has already proven by reducing infections, hospitalisations and mortality due to COVID-19 [4,5]. Therefore reduced immunization against COVID-19 after a few months of vaccination makes booster vaccination necessary especially for vulnerable groups. However, a meta-analyis found that adverse effects and discomfort experienced after primary COVID-19 vaccination were the most important reasons for decline of the first COVID-19 booster dose [6].

Unfortunately, despite the high safety and efficacy of most COVID-19 vaccines, there are also adverse effects. The most common adverse effects are fever, fatigue, muscle aches, pain and swelling at the injection site, menstrual disorders and headache, while more serious adverse effects such as myocarditis, pericarditis and thrombosis are extremely rare [7–10]. Other minor adverse effects that occur less frequently are nausea, shortness of breath, and dizziness. In particular, a systematic review with 14 studies and 10,632 participants found that 77.3% developed pain at the injection site, 43% felt a shooting pain, 39.7% developed muscle pain, 34% developed swelling at the injection site, 33.3% developed headaches, 18.3% developed chills, 18% developed fever, 8% developed swollen lymph nodes, 7.9% developed nausea, 7.6% developed shortness of breath, and 6.4% developed diarrhea [11]. Another systematic review investigated menstrual abnormalities and found that more than half of females (52.1%) had some form of a menstrual problem after COVID-19 vaccines (e.g., μenorrhagia, metrorrhagia, and polymenorrhea) in a total of 78,138 vaccinated females [12]. Younger people and females are more likely to experience adverse effects [13]. For example, a systematic review found that 69.8% of females developed adverse effects, while the corresponding percentage for males was 30.2% [11].

To date, few studies have investigated adverse effects after the first COVID-19 booster dose focusing only on demographic variables such as gender and age as possible risk factors [14–19]. Adverse effects after the first COVID-19 booster dose are similar with those after the primary doses and are analyzed above.

However, to the best of our knowledge, there are no studies that assess adverse effects after COVID-19 vaccination in Greece. Thus, the aim of our study was to estimate the prevalence of adverse effects after the first COVID-19 booster dose in Greece, and to identify possible risk factors.

## Material and methods

### Study design

We conducted a cross-sectional study in Greece. Data collection was performed during November 2022. Inclusion criteria were adults over 18 years old, individuals that understand the Greek language, and individuals that have been vaccinated against SARS-CoV-2 with the primary COVID-19 vaccine doses and the first booster dose. Our participants visited a primary health center in Athens to receive a second booster dose. Thus, a convenience sample from the general population was obtained. Response rate was 94.6% (473 out of 500).

We measured the following adverse effects: pain at the injection site, swelling at the injection site, fatigue, muscle pain, headaches, fever, chills, nausea, itching, diarrhea, shortness of breath, rhinorrhea, swollen lymph nodes, dizziness, sleep disturbances, reduced appetite, and adverse effects on menstrual cycle. Also, we asked participants to add any other possible adverse effect that experienced because of vaccination but it was not included in our questionnaire. Each adverse effect was measured on a four-point scale: not at all effect, minor effects, moderate effects, and major effects. Moreover, we added all adverse effects in order to calculate a total score of adverse effects.

We considered gender (males or females), age (continuous variable), chronic disease (no or yes), self-assessment of health status (very poor, poor, moderate, good, very good), COVID-19 diagnosis (no or yes), and self-assessment of COVID-19 clinical course (scale from 0 [very mild] to 10 [extreme severe] as possible predictors of frequency of adverse effects of COVID-19 booster dose. Moreover, we considered a priory that scores of COVID-19 clinical course from 0 to 2 indicate low symptoms of COVID-19, scores from 3 to 7 indicate moderate symptoms, and scores from 8 to 10 indicate extreme symptoms.

Taken into our consideration that the reference population of fully vaccinated adults in Greece was 7.7 million at the time of our study, a confidence level of 95%, a margin of error of 5%, and a population proportion of 50%, the minimum sample size was 385 participants. We decided to increase slightly our sample in order to decrease random error.

### Ethics

We informed the participants about the aim and the design of our study and they gave their informed consent to participate in our study. We did not collect personal data of the participants. Thus, participation in our study was anonymous and voluntary.

Guidelines of the Declaration of Helsinki were applied in our study. Also, our study protocol was approved by the Ethics Committee of Faculty of Nursing, National and Kapodistrian University of Athens (reference number; 424, 26-10-2022).

### Statistical analysis

We present categorical variables with numbers and percentages. Also, we present continuous variables with mean and standard deviation. Kolmogorov-Smirnov test and Q-Q plots indicated that continuous variables followed normal distribution. We considered gender, age, chronic disease, self-assessment of health status, COVID-19 diagnosis, and self-assessment of COVID-19 clinical course as independent variables, and adverse effects of COVID-19 booster dose as dependent variables. We used chi-square test, independent samples t-test, Pearson’s correlation coefficient, and Spearman’s correlation coefficient to assess the relationship between independent variables and adverse effects. P-values less than 0.05 were considered as statistically significant. We used the IBM SPSS 21.0 (IBM Corp. Released 2012. IBM SPSS Statistics for Windows, Version 21.0. Armonk, NY: IBM Corp.) for the analysis.

## Results

Study population included 473 participants. Demographic characteristics of participants are shown in Table 1. Mean age of participants was 38.4 years, while most of them were females (78.6%). Among them, 19.2% had a chronic disease. The majority of participants reported that their health status was good/very good (91.7%), while 5.3% reported a moderate level of health, and 2.9% a poor/very poor level of health.

**Table 1.** Demographic and COVID-19-related variables of participants (N=473).

| Variables | N | % |
| --- | --- | --- |
| Gender |  |  |
| Males | 101 | 21.4 |
| Females | 372 | 78.6 |
| Age (years), mean, standard deviation | 38.4 | 9.9 |
| Chronic disease |  |  |
| No | 382 | 80.8 |
| Yes | 91 | 19.2 |
| Self-assessment of health status |  |  |
| Very poor | 13 | 2.7 |
| Poor | 1 | 0.2 |
| Moderate | 25 | 5.3 |
| Good | 274 | 57.9 |
| Very good | 160 | 33.8 |
| COVID-19 diagnosis |  |  |
| No | 145 | 30.7 |
| Yes | 328 | 69.3 |
| COVID-19 symptoms |  |  |
| Low | 110 | 33.5 |
| Moderate | 203 | 61.9 |
| Extreme | 15 | 4.6 |

In our sample, 69.3% have been diagnosed with COVID-19 during the pandemic (Table 1). Among them, 61.9% developed moderate symptoms of COVID-19, 33.5% developed low symptoms, and 4.6% developed extreme symptoms.

Adverse effects of COVID-19 booster dose in participants are shown in detail in Table 2. In our sample, 96% (n=454) had at least one adverse effect. Half of the participants (50.2%, n=239) had one to five adverse effects, 35.9% (n=170) had six to ten adverse effects, and 9.5% (n=45) had 11 to 16 adverse effects. Mean number of adverse effects was 5.5 (standard deviation = 3.5) with a median value of 5, a minimum value of 0 and a maximum value of 16.

**Table 2.** Adverse effects of COVID-19 booster dose in participants.

| Adverse effects | Not at all |  | Minor |  | Moderate |  | Major |  |
| --- | --- | --- | --- | --- | --- | --- | --- | --- |
|  | N | % | N | % | N | % | N | % |
| Pain at the injection site | 74 | 15.6 | 178 | 37.6 | 152 | 32.1 | 69 | 14.6 |
| Swelling at the injection site | 212 | 44.8 | 147 | 31.1 | 80 | 16.9 | 34 | 7.2 |
| Fatigue | 138 | 29.2 | 121 | 25.6 | 106 | 22.4 | 108 | 22.8 |
| Muscle pain | 184 | 38.9 | 109 | 23.0 | 89 | 18.8 | 91 | 19.2 |
| Headache | 237 | 50.1 | 89 | 18.8 | 91 | 19.2 | 56 | 11.8 |
| Fever | 270 | 57.1 | 87 | 18.4 | 69 | 14.6 | 47 | 9.9 |
| Chills | 279 | 59.0 | 92 | 19.5 | 47 | 9.9 | 55 | 11.6 |
| Nausea | 413 | 87.3 | 30 | 6.3 | 16 | 3.4 | 14 | 3.0 |
| Itching | 347 | 73.4 | 90 | 19.0 | 26 | 5.5 | 10 | 2.1 |
| Diarrhea | 450 | 95.1 | 10 | 2.1 | 11 | 2.3 | 2 | 0.4 |
| Shortness of breath | 440 | 93.0 | 23 | 4.9 | 6 | 1.3 | 4 | 0.8 |
| Rhinorrhea | 412 | 87.1 | 38 | 8.0 | 20 | 4.2 | 3 | 0.6 |
| Swollen lymph nodes | 348 | 73.6 | 53 | 11.2 | 38 | 8.0 | 34 | 7.2 |
| Dizziness | 371 | 78.4 | 55 | 11.6 | 31 | 6.6 | 16 | 3.4 |
| Sleep disturbances | 406 | 85.8 | 36 | 7.6 | 18 | 3.8 | 13 | 2.7 |
| Reduced appetite | 392 | 82.9 | 48 | 10.1 | 17 | 3.6 | 16 | 3.4 |
| Menstrual cycle | 281 | 75.5 | 36 | 9.7 | 28 | 7.5 | 27 | 7.3 |

The most frequent adverse effects were pain at the injection site (84.3%), fatigue (70.8%), muscle pain (61%), swelling at the injection site (55.2%), headache (49.8%), fever (42.9%), and chills (41%). Itching (26.6%), swollen lymph nodes (26.4%), adverse effects on menstrual period (24.5%), dizziness (21.6%), and reduced appetite (17.1%) were frequent adverse effects. Moreover, sleep disturbances (14.1%), rhinorrhea (12.8%), nausea (12.7%), shortness of breath (7%), and diarrhea (4.8%) were rare adverse effects.

Relationships between demographic and COVID-19-related variables and total number of adverse effects of COVID-19 booster dose are shown in Table 3. Females developed more adverse effects than males (p<0.001). Also, we found a positive relationship between severity of COVID-19 symptoms and adverse effects of COVID-19 booster dose (p=0.005). Moreover, decreased age was associated with increased adverse effects (p<0.001).

**Table 3.**
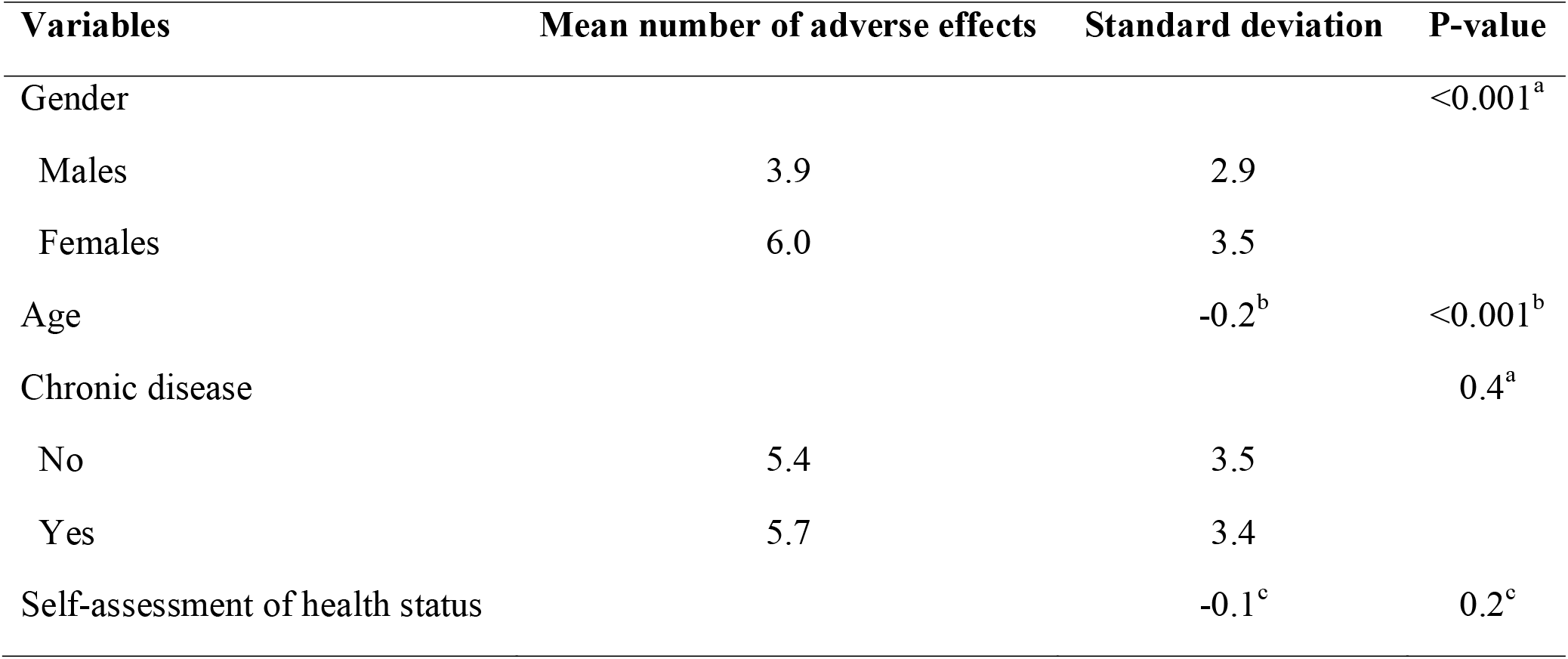
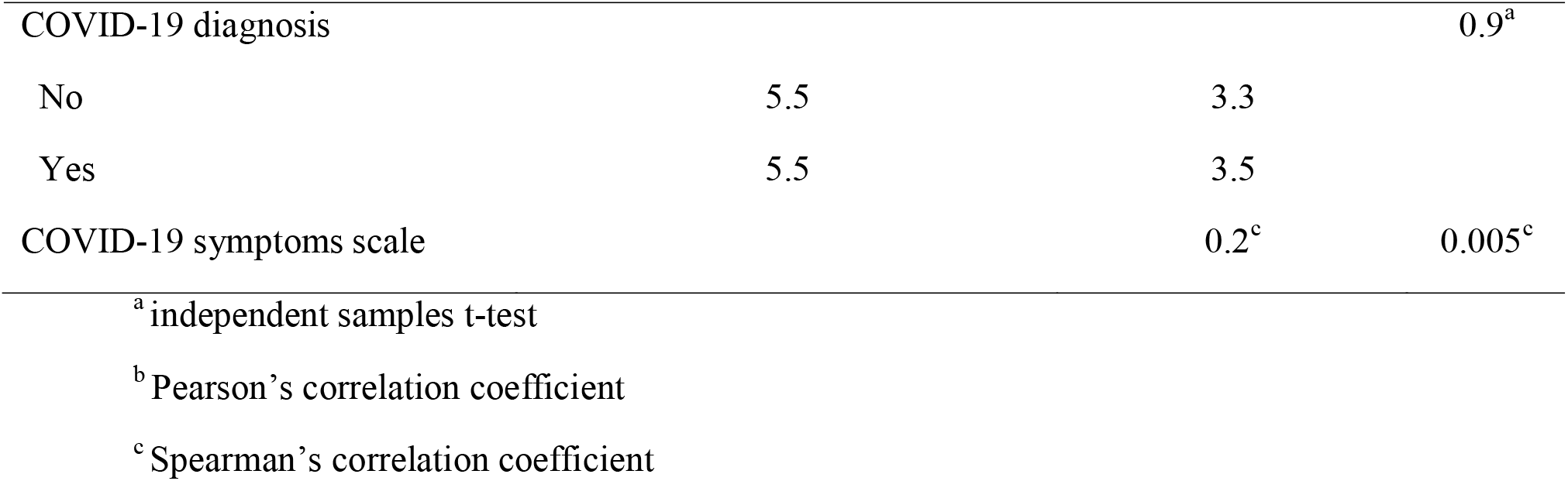
Relationships between demographic and COVID-19-related variables and total number of adverse effects of COVID-19 booster dose.

| Variables | Mean number of adverse effects | Standard deviation | P-value |
| --- | --- | --- | --- |
| Gender | | | $<0.001^a$ |
| Males | 3.9 | 2.9 |  |
| Females | 6.0 | 3.5 |  |
| Age | | $-0.2^b$ | $<0.001^b$ |
| Chronic disease | | | $0.4^a$ |
| No | 5.4 | 3.5 |  |
| Yes | 5.7 | 3.4 |  |
| Self-assessment of health status | | $-0.1^c$ | $0.2^c$ |
| COVID-19 diagnosis |  |  | 0.9<sup>a</sup> |
| No | 5.5 | 3.3 |  |
| Yes | 5.5 | 3.5 |  |
| COVID-19 symptoms scale |  | 0.2<sup>c</sup> | 0.005<sup>c</sup> |

<sup>a</sup> independent samples t-test
<sup>b</sup> Pearson's correlation coefficient
<sup>c</sup> Spearman's correlation coefficient

We present statistically significant relationships between demographic and COVID-19-related variables and adverse effects separately in Table 4. Females developed more often pain at the injection site (p=0.011), swelling at the injection site (p<0.001), fatigue (p=0.009), muscle pain (p<0.001), headache (p<0.001), chills (p=0.017), itching (p<0.001), rhinorrhea (p=0.019), swollen lymph nodes (p<0.001), dizziness (p=0.003), and sleep disturbances (p=0.008). Younger participants developed more often muscle pain (p=0.002), headache (p=0.012), fever (p<0.001), chills (p<0.001), shortness of breath (p=0.003), swollen lymph nodes (p=0.008), dizziness (p=0.004), and adverse effects on menstrual cycle (p<0.001). Participants with worse COVID-19 clinical course had more often headache (p=0.003), fever (p=0.021), rhinorrhea (p=0.014), and adverse effects on menstrual cycle (p=0.001).

**Table 4.** Statistically significant relationships between demographic and COVID-19-related variables and adverse effects of COVID-19 booster dose.

| Adverse effects | Gender<sup>a</sup> |  | P-value<sup>b</sup> | Age<sup>c</sup> | P-value<sup>d</sup> | COVID-19 symptoms scale<sup>c</sup> | P-value<sup>d</sup> |
| --- | --- | --- | --- | --- | --- | --- | --- |
|  | Males | Females |  |  |  |  |  |
| Pain at the injection site | 77 (76.2) | 322 (86.6) | 0.011 |  |  |  |  |
| Swelling at the injection site | 39 (38.6) | 222 (59.7) | <0.001 |  |  |  |  |
| Fatigue | 61 (60.4) | 274 (73.7) | 0.009 |  |  |  |  |
| Muscle pain | 42 (41.6) | 247 (66.4) | <0.001 |  | 0.002 |  |  |
| No |  |  |  | 40.3 (11.1) |  |  |  |
| Yes |  |  |  | 37.2 (8.9) |  |  |  |
| Headache | 32 (31.7) | 204 (54.8) | <0.001 |  | 0.012 |  | 0.003 |
| No |  |  |  | 39.6 (10.4) |  | 3.3 (2.3) |  |
| Yes |  |  |  | 27.3 (9.2) |  | 4.1 (2.2) |  |
| Chills | 31 (30.7) | 163 (43.8) | 0.017 |  | <0.001 |  |  |
| No |  |  |  | 39.7 (10.6) |  |  |  |
| Yes |  |  |  | 36.6 (8.4) |  |  |  |
| Itching | 11 (10.9) | 115 (30.9) | <0.001 |  |  |  |  |
| Rhinorrhea | 6 (5.9) | 55 (14.8) | 0.019 |  |  |  | 0.014 |
| No |  |  |  |  |  | 3.6 (2.3) |  |
| Yes |  |  |  |  |  | 4.5 (2.3) |  |
| Swollen lymph nodes | 9 (8.9) | 116 (31.2) | <0.001 |  | 0.008 |  |  |
| No |  |  |  | 39.2 (10.1) |  |  |  |
| Yes |  |  |  | 36.4 (8.9) |  |  |  |
| Dizziness | 11 (10.9) | 91 (24.5) | 0.003 |  | 0.004 |  |  |
| No |  |  |  | 39.1 (10.2) |  |  |  |
| Yes |  |  |  | 36.2 (8.4) |  |  |  |
| Sleep disturbances | 6 (5.9) | 61 (16.4) | 0.008 |  |  |  |  |
| Fever |  |  |  |  | <0.001 |  | 0.021 |
| No |  |  |  | 39.9 (9.9) |  | 3.5 (2.2) |  |
| Yes |  |  |  | 36.5 (9.6) |  | 4.1 (2.4) |  |
| Shortness of breath |  |  |  |  | 0.003 |  |  |
| No |  |  |  | 38.8 (9.9) |  |  |  |
| Yes |  |  |  | 33.5 (8.9) |  |  |  |
| Menstrual cycle |  |  |  |  | <0.001 |  | 0.001 |
| No |  |  |  | 39.0 (9.8) |  | 3.5 (2.2) |  |
| Yes |  |  |  | 35.3 (7.2) |  | 4.6 (2.4) |  |

<sup>a</sup> number (percentage)
<sup>b</sup> chi-square test
<sup>c</sup> mean (standard deviation)
<sup>d</sup> independent samples t-test

## Discussion

We conducted a cross-sectional study in Greece to assess the prevalence of adverse effects after COVID-19 vaccines, and to investigate possible risk factors. To the best of our knowledge this is the first study that estimates adverse effects after COVID-19 vaccination in Greece.

We found that almost all participants (96%) developed at least one adverse effect after the first booster dose. Literature supports this finding since two other studies found that all participants had adverse effects after the first booster dose [20,21]. Other studies found also high prevalence of adverse effect after the first booster dose ranging from 74.7% to 89% [14–17,22,23]. Slight differences in prevalence of adverse effects among studies could be attributed to different study designs, populations, and definitions.

Our study showed that the most frequent adverse effects were pain at the injection site, fatigue, muscle pain, swelling at the injection site, headache, fever and chills. These findings are in accordance with the literature regarding the adverse effects of the first booster dose [14–18,22,23]. Moreover, a recent systematic review investigated the prevalence of adverse effects after the primary COVID-19 vaccine doses and found that the most frequent adverse effects among 10,632 participants were pain at the injection site (77.3%), muscle pain (39.7%), swelling at the injection site (34%), headache (33.3%), chills (18.3%), fever (18%), swollen lymph nodes (8%), nausea (7.9%), and shortness of breath (7.6%) [11]. Fortunately, although adverse effects after COVID-19 vaccination are common, they are usually mild and self-limited [11].

Also, we found a positive relationship between severity of COVID-19 symptoms and adverse effects of COVID-19 booster dose. To our knowledge, our study investigated for first time the relationship between severity of COVID-19 symptoms and adverse effects and thus a direct comparison with other studies could not be feasible. However, it is well known that adverse effects are more common among individuals with a history of COVID-19 or/and past exposure to SARS-CoV-2 not only after the primary vaccination [24–26] but also after the first booster dose [16,20]. Repeated COVID-19 vaccine doses can induce more adverse effects due to cytokine production especially in people who had previously been infected with SARS-CoV-2 [27]. Moreover, humoral immunity in people with a history of COVID-19 and a single dose of the COVID-19 vaccine is equal to or stronger than in uninfected individuals with two doses [28].

According to our results females developed more adverse effects than males. Several studies confirm the fact that adverse effects are more common among females after the first booster dose [14,16,17,20]. Several theories could explain this gender-based difference in adverse effects after COVID-19 vaccination. Firstly, in general, females have a greater response to vaccines than males due to antibody, inflammatory, and cell-mediated immune responses [29,30]. Additionally, genetic, hormonal, and behavioral factors might also contribute to the explanation of the higher frequency of adverse events in females [30–33]. For example, the innate and adaptive immune response in males has been depressed by testosterone [34,35]. Since early evidence shows that females are less willing to receive a second COVID-19 booster dose [36], high incidence of adverse effects among females could decrease their willingness to accept a booster dose even more.

Moreover, we found a negative relationship between age and incidence of adverse effects after the first booster dose. Several studies confirm our finding since they revealed that younger adults were more likely to develop adverse effects after COVID-19 vaccination compared to older adults [37–39]. Adverse effects after COVID-19 vaccination are a by-product of the exuberant production of type-I interferon that happens to start an effective immune response against the SARS-CoV-2 [40]. Since the generation of type-I interferon is more potent in younger adults this mechanism can explain the higher prevalence of adverse effects among them [40,41].

Our study had several limitations. First, although we achieved the minimum required sample size we obtained a convenience sample from a single center. Therefore, our sample might not be representative of the vaccinated adults in Greece. For example, our participants were predominantly young females. Further studies with bigger and more representative samples could add valuable information. Second, we used a self-reported questionnaire to collect our data. Since we cannot perform a clinical confirmation an information bias is possible in our study. Additionally, a recall bias is possible since our participants have been received the booster dose at least six months before our study. Timely clinical evaluation of individuals after COVID-19 vaccination in future studies could minimize information and recall bias. Third, we performed a cross-sectional study and therefore we cannot establish a causal relationship between the risk factors and incidence of adverse effects. Fourth, we investigated several risk factors of adverse effects but future research should expand our knowledge on this field. Fifth, since we performed our study in a single center all participants received the same COVID-19 vaccine and thus comparisons between different vaccines cannot be made. Sixth, we only measured short-term adverse effects without assessing the duration of these effects. Finally, we did not measure the medical status of participants (e.g. comorbidities) and the way this could have influenced the adverse effects of the booster dose.

## Conclusions

Almost all participants in our study developed adverse effects after the first booster dose. However, these adverse effects were minor and not life-threatening. Moreover, our results showed that female gender, worse clinical course of COVID-19, and decreased age were associated with an increased number of adverse effects. Our study provides important baseline data for policy makers, healthcare professionals, and the general public to be aware of adverse effects after booster doses.

COVID-19 vaccines are the most effective weapon to reduce hospitalizations, intensive care admissions, and deaths from the disease. Booster doses are offered now seasonally and they are highly recommended for vulnerable groups such as immunosuppressed people. Therefore, monitoring of adverse effects of booster doses is essential in order to increase public confidence in the safety profile of vaccines and improve vaccine coverage in high-risk groups. Moreover, identification of risk factors is crucial to protect individuals’ and communities’ health.

General public’s awareness of adverse effects could reduce hesitancy and convince vulnerable people to accept booster doses.

## Data Availability

All data produced in the present study are available upon reasonable request to the authors

